# Effectiveness of continuous Ketamine infusion associated with magnesium sulfate for management of patient with chronic pain: A prospective observational study (Kontinue)

**DOI:** 10.1101/2025.06.01.25328713

**Authors:** Amodeo Jean-Marie, Berthier Frederic

## Abstract

**Background:** Chronic pain is usually refractory to treatment. Ketamine is often used to treat chronic refractory pain.In France most of the patients are treated in pain units day hospital.

This study aims to evaluate the effectiveness of continuous four days of Ketamine infusion associated with magnesium sulfate for inpatients.

**Methods:** A total of 89 patients received a 4 days continuous subanesthetic IV ketamine infusion associated with 1000 mg/day of magnesium sulfate.Primary outcome was mean pain intensity (Numerical Rating Pain Scale) 30 days after infusion. Patient Impression of Change (PGIC), Anxiety and depression (HADS), Neuropathic Pain (NPSI), quality of life (SF-12) were secondary outcomes.

**Results:** Ketamine continuous infusion was associed with decrease of NRS (7.54 ± 1.35 at d0 to 5.59 ± 2.2 at d30 [26%; p < 0.001]).The PGIC improved in 68.7 % patients (36.2 % reports “much improved” or “very much improved”).There was a significant decrease of Npsi total score (−28% ; p < 0.0001) and an increase of SF12 (Mental health dimension from 31.4 ±7.9 to 36.4 ± 8.9 [+15.9 % ; p < 0.0001]; Physical health dimension from 30.2 ± 6.7 to 32.8 ± 6.8 [+8.6% ; p:0.0007]).The mean HADS Depression decreased from 10 ± 4.5 at d0 to 8.2 ± 4.6 at d30 (−15% ; p:0.0012) ; The mean HADS Anxiety decreased from 11.35 ± 4.7 at d0 to 9.38 ± 4.5 at d30 (−14% ; p < 0.0001).

Conclusions Findings suggests that continuous low-dose infusion of ketamine is effective for chronic pain relief to inpatients.

## Introduction

The management of chronic pain presents a worldwide major challenge for healthcare professionals, it require an multidisciplinary approach to care the components of the patient’s pain.

The prevalence of chronic pain conditions approaches 20% worldwide (1). It impacts quality of life significantly (2) and prevalence rates for pain are expected to increase as populations continue to age (3).

Development of a treatment plan includes establishing a diagnosis, and measurable outcomes that focus on improvements such as quality of life (4).

The multimodal therapeutic approach includes both pharmacological and non-pharmacological modalities for pain.

In chronic pain states the NMDAR is activated and upregulated in the spinal cord (central sensitization), resulting in enhanced signal transmission in the pain from the spinal cord to the cortex leading to spontaneous pain, allodynia and hyperalgesia (5) (6).

Ketamine is regularly used in the management of chronic pain patients.

In recent years, numerous studies have been published on ketamine for the treatment of chronic non-cancer pain with different dosages, routes of administration, infusion durations (7)(8), with controversial results and level of evidence varies by condition and dose range (9).Several studies in France have been carried out (10,11,12) in order to clarify the practical methods of use and effective dosages.

Concerning the routes of administration, a group of experts noted intravenous infusion at doses of 0.5 to 0.9 mg/kg/day for four days of treatment as preferable (13). Most of ketamine infusions in France are delivered in Pain units day hospital.

The objective of the Kontinue study is to evaluate the effectiveness of continuous infusion of ketamine 0.5 mg mg/kg/24h for 4 days combined with magnesium sulfate for the management of patients suffering from chronic pain.

## Materials and Methods

### Study design and population

This is a prospective single-center observational study based on the collection of validated self-questionnaires: Numeric rating pain scale (NRS), Patient Global Impression of Change (CGI-C), Neuropathic Pain Symptom Inventory (NPSI), Hospital Anxiety and Depression Scale (HAD), Short Form 12 (SF12), carried out at the Princess Grace Hospital Center in Monaco. Registred in ClinicalTrials ID : NCT06131970.The study was conducted in accordance with the Declaration of Helsinki, approved by the local Ethics Committee, and written informed consent was obtained from all participants before procedure.

The inclusion criteria were as follow: Pain lasting for more than six months; Monitoring in CHPG Monaco; had a psycho-social assessment and have been the subject of multidisciplinary discussion recorded in the medical record ; to be volunteer to participate in the study. Exclusion criteria included Inability to give consent; without contraindication to treatment.

The objective was to determine the effectiveness on pain at one month of Ketamine injection in continuous intravenous infusion for 4 days with a dose of 0.5 mg/kg/day, combined with magnesium sulfate 1000 mg/day.

Recommendations are consistent regarding core domains which should be assessed in pain clinical trials, including: pain (intensity, quality), physical functioning (daily activities/well-being), emotional functioning, and patient global improvement and satisfaction with treatment (14).

The data collected concerns 89 patients; the main criteria was pain intensity (0-10 numerical pain rating scale) at one month. The secondary endpoints were the improvement of global impression of change (CGI-C), neuropathic pain (NPSI:The quality of neuropathic pain was categorized in 5 sub-groups: superficial, deep, paroxysmal, evoked, and dysaesthetic : each rated on a numeric scale from 0 to 10), the benefit on quality of life and mood (SF12;HAD) at day 30; the number of patients still improved at 6 moths was collected. (figure 1)

**Figure 1.**
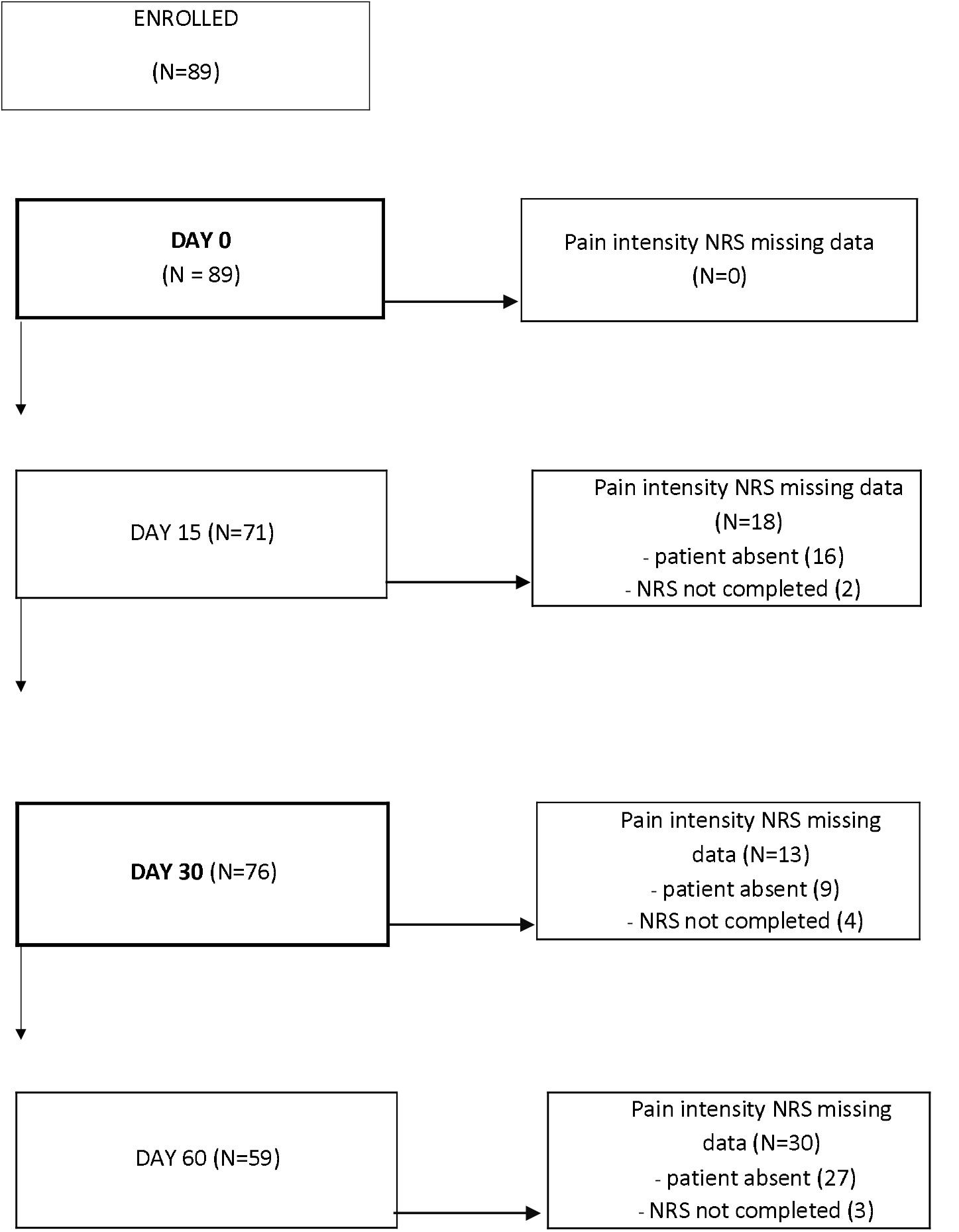
Flow chart of the study. NRS Numeric rating scale, NRS at DAY 30 = primary endpoint

### Statistical analysis

Descriptive statistics used mean and standard deviation for continuous variables and the number of cases and percentages for categorical variables. Pain intensity NRS improvement was assessed by Wilcoxon signed-rank test. Between groups comparisons were performed with the Chi-squared test for categorical variables and the Wilcoxon Rank Sum Test for continuous variables. Spearman’s rank correlation coefficients were used to estimate the relationship between scales. P-values ≤ 0.05 were considered significant. Statistical analyses were performed using SAS 9.1 (SAS Institute, Cary, NC, USA).

## Results

### Patient baseline characteristics

Of a total of 89 patients were included in this study 87.6% were female and 12.4% were male. The mean age was 52.2 ± 9.6 years (Min 20 to Max 74). The mean NRS was 7.54 ± 1.35. We found three principal etiology of pain: 59.5 % had fibromyalgia, 33.7 % of Neuropathic pain (without fibromyalgia) ; 6.6 % had arthosis pain (without neuropathic pain). The others results concerning baseline score are presented in table 1

**Table 1:**
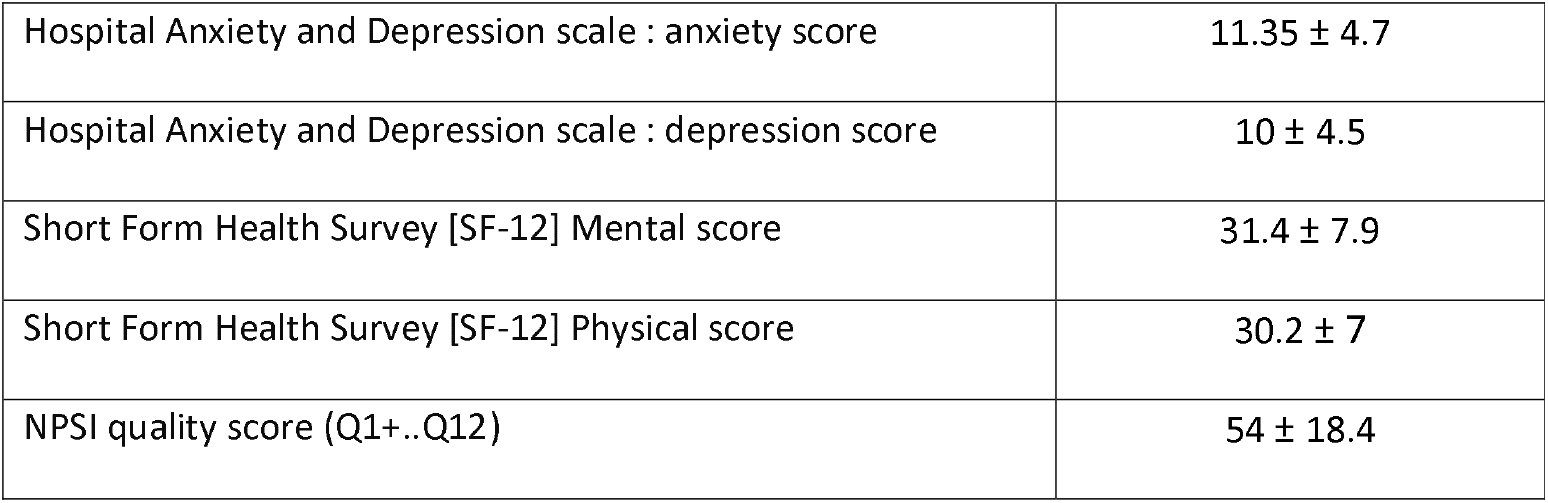
Baseline HADS (AD) ; SF12 (MCS, PCS); NPSI.

### Effect of treatment

#### Main criteria: Pain Intensity (NRS)

#### NRS

NRTSh e intensity of pain improved during the observation period: the average NRS score decreased from 7.54 ± 1.35 at d0 to 5.59 ± 2.2 at d30 (26% ; p<0.001). NRS score at d60 was 5.29 ± 2.5 (table 2). Pain relief was correlated with diminution of anxiety (Spearman coefficient 0.33 ; p:0.0044) and depression (Spearman coefficient 0.41 ; p:0.0004).

**Table 2:**
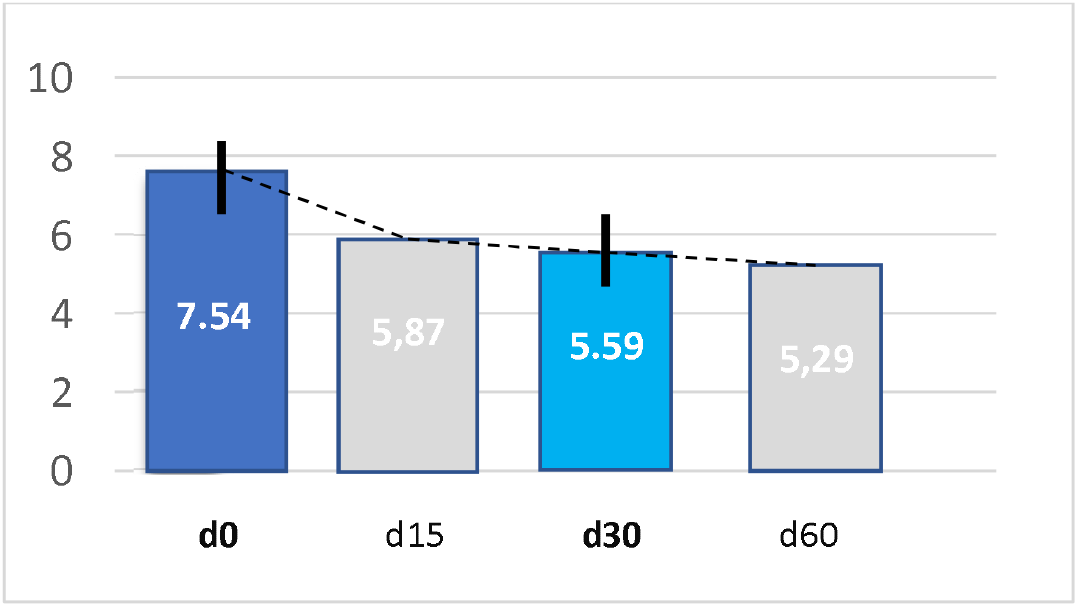
Pain intensity.

Patient Global Impression of Change (PGIC): The improvement was observed in 68.7 % patients (n :80) and 36.2 % reports “much improved” or “very much improved” impression of change at day30. The PGIC was correllated with improvement of NRS score : -2.9 ± 2 (very much improved : -4.5 ± 1.7 ; much improved : -3.5 ± 2.1 ; improved : - 1.88 ± 1.39).

In the improved patients group we noticed a significant diminution of HAD-anxiety (−2.2 ± 2.9) and HAD-depression (−2.19 ± 3.9).

Changes from baseline in HAD and PGIC groups are reported in table 3 and 4.

**Table 3.**
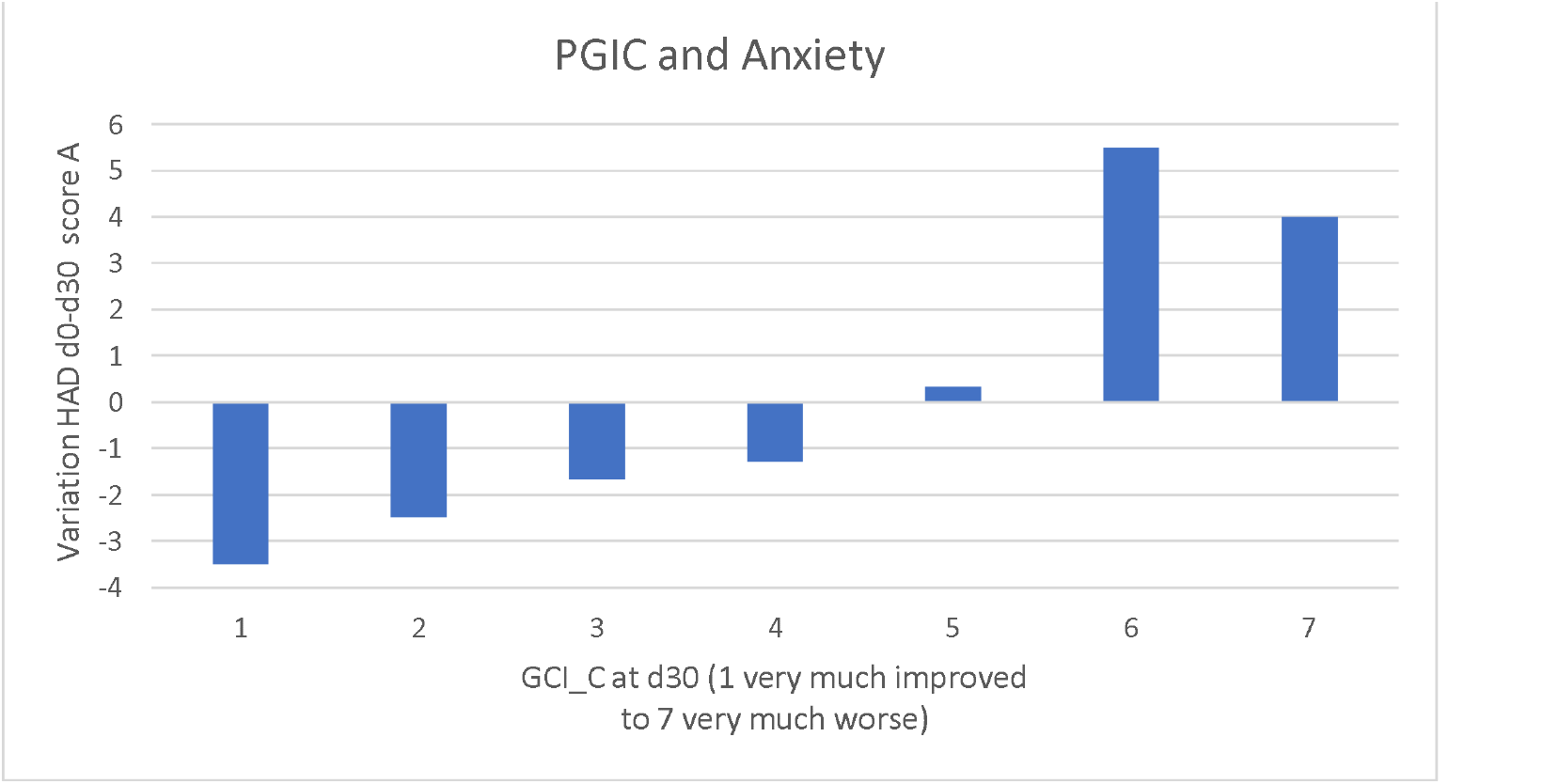
Variation of HAD anxiety score in all groups of PGIC.

**Table 4.**
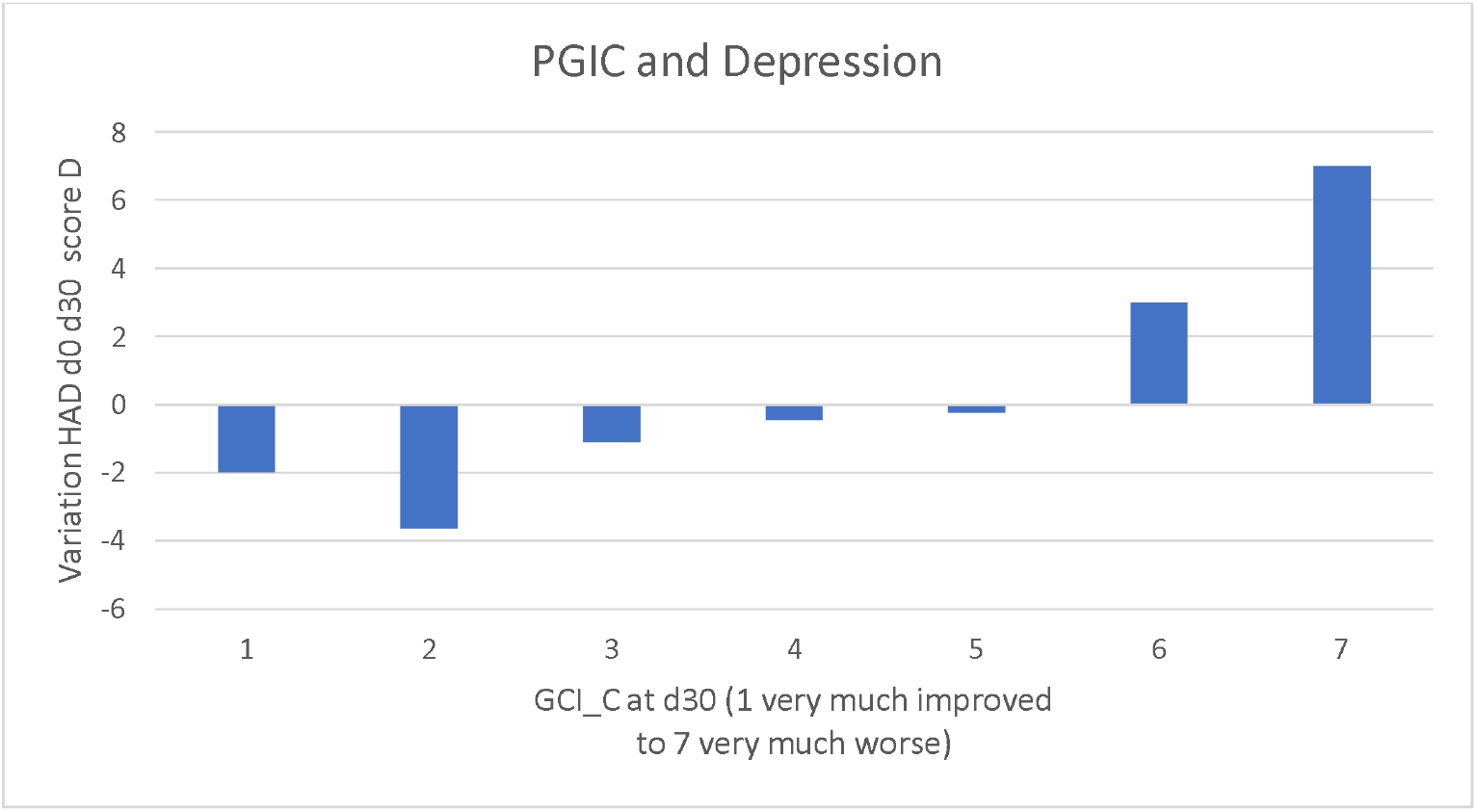
Variation of HAD depression score in all groups of PGIC.

The mean HADS Depression decreased from 10 ±4.5 at d0 to 8.2 ± 4.6 at d30 (−15% ; p=0.0012) ; The mean HADS Anxiety decreased from 11.35 ±4.7 at d0 to 9.38 ±4.5 at d30 (−14% ; p<0.0001). The number of patients with HAD Anxiety status≥ 11 at d0 decreased of 12.2% at d30 (from 53% to 41.8 %) and -17.5% for depression score ≥ 11 (44% to 28.2 %).

Patients with HADS anxiety score <11 had better improvement NRS compared with patients HADS score ≥ 11 : -2.62 vs -1.44 (p:0.04).

The mean Mental health dimension score of SF12 increased from baseline to d30, from 31.4 ±7.9 to 36.4 ± 8.9 (+15.9 % ; p<0.0001); The mean physical health dimension score of SF12 increased from 30.2 ± 7.0 to 32.8 ± 6.9 (+8.6% ; p:0.0007). Analysis showed MCS was strongly associated with pain diminution (coefficient - 0.41; p:0.0007) ; PCS improvement was associated with NRS (coefficient -0.45 ; p:0.0001).

The intensity of the neuropathic pain evaluated by NPSI, showed an improvement from baseline (NPSI total score) 54 ± 18.4 to 38.4 ± 22.4 at d30 (−28% ; p<0.0001). Total score corresponding to the sum of the scores of the 10 sensory descriptors. All the sub groups were statistically and significantly decreasing from baseline (table 5).

**Table 5:**
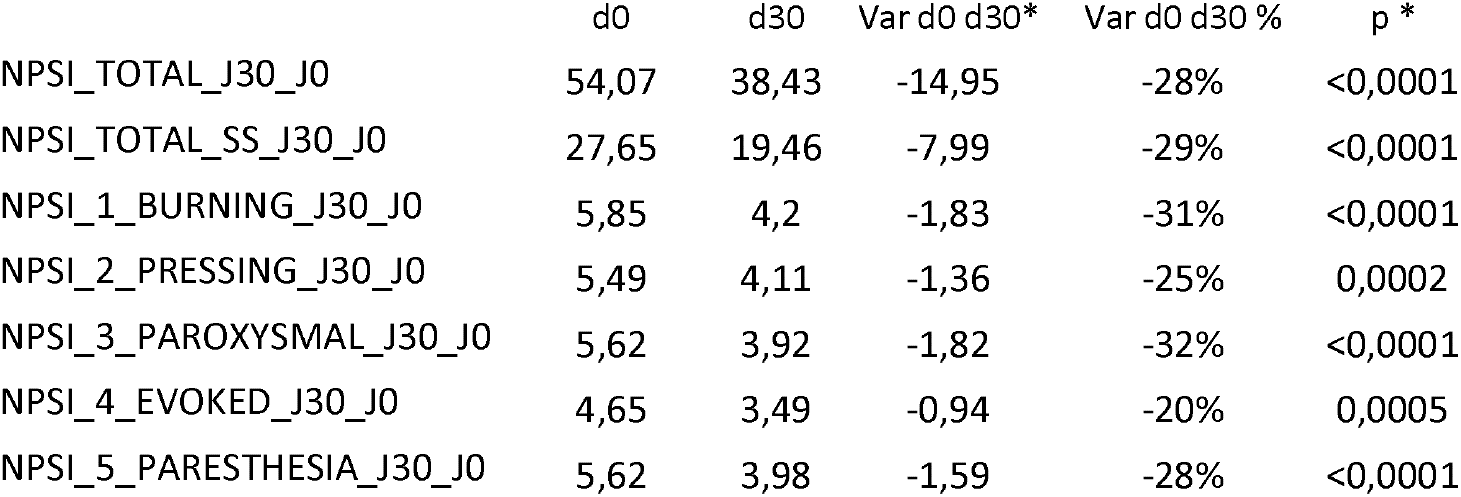
NPSI total and sub-groups evolution d0 to d30. ^*^patients without missing data at d0 and d30 (59 for total ; 77 for burning and paresthesia ; 70 for pressing ; 76 for paroxysmal ; 67 for evoked)

The mean duration of pain relief was 3 months (2.99 ± 2.29 [3.4 ± 2.2 for fibromyalgia and 2.6 ± 2.2 for neuropathic pain]) ; The percentage of patients still improved at 6 months seemed to be higher in fibromyalgia patients but not significatively (p:0.18).

Supplemental table 1 compares patients according to missing data for main criteria at day 30. Both population do not differ for age, sex, pain intensity, SF-12 mental and physical scores and HAD anxiety score. Patients with missing data had less fibromyalgia and higher HAD anxiety score (p = 0.04):

## Discussion

Our results revealed safety and efficiency of low dose and slow intravenous infusion of ketamine.

This observational study showed that 4 days of low doses (0.5 mg/kg/d) continuous ketamine infusion for an inpatient, associed with Magnesium Sulfate, is an effective treatment for chronic pain.

Usually the protocol and infusion rate belong to clinical experience but also to availability of hospitalization type (outpatient,inpatient). We used a 24 h infusion inpatient for best tolerance and longer lasting analgesia. There is evidence that the duration of infusion determines the duration of analgesic effect (15). We found also at d60 an improvement of pain but with data collected on only 59 patients (NRS 5.29 ±2.5).

We did not observe any serious adverse events associated with continuous infusion of ketamine. None required to stop the treatment

In several studies, the total dose of ketamine was higher with long pain relief and also sides effects (16,17).

Patients received Magnesium sulfate according to the additive effect with ketamine because both molecules share the NMDA receptor as a target of action (18).

Our study therefore includes patients with neuropathic pain as well as with fibromyalgia (fulfilling the American College of Rheumatology classification criteria) and local arthosis. Ketamine use has typically been strongest in neuropathic pain. In subgroup analysis, there is no evidence for superior analgesic benefit from ketamine in neuropathic compared to nociceptive or nociplastic pain syndromes (19).

All the data collected with common pain questionnaires showed improvement at day 30. The PGIC improvement was correlated with NRS improvement and HADS.

The response to the treatment was similar in the 5 dimensions of neuropathic pain.

In the results it must be taken into account that there is neuropathic pain in fibromyalgia. The mean duration of pain relief was 3 months (2.99 ± 2.29) and 22 % of patients had no recurrence of pain at 6 months. The number of placebo-controlled randomized controlled trials (RCTs) on long term ketamine treatment (from 4 days) is limited but indicating that the effect of ketamine treatment can persists for at least 41weeks (20).

### Limitations

This stydy is limited by its observationnal nature,as well as the single center design. Additionnaly the absence of placebo group is a limitation, the placebo effect of ketamine has not been evaluated. We didn’t report results at d60 regarding to insufficient collected data (70% of patients responding to outcome data PGIC and HAD).Although the sample size of 89 patients may limit the generalizability of our results.

Therefore any changes to pain scores may have been influenced by patients : for exemple some patients reporting improvement of global quality of life had stopped there usual treatment like antiepileptics causing reccurence of pain.

We reported an additional table comparing patients with and without missing data for the main endpoint: the risk of bias is limited as initial NRS levels are comparable.

There is a growing body of evidence demonstrating analgesic effects in inflammatory pain conditions (21,22) which have not been evaluated in our findings.

Therefore, future studies should involve large patient samples with more long-term treatment and follow up, and placebo-controlled prospective trial are needed to confirm efficiency of low and slow dose of ketamine.

Nevertheless, this study showed significant reduction of NRS in refractory chronic pain patients following 4 days of IV ketamine infusion associated with daily IV magnesium sulfate.

## Conclusion

The Kontinue study showed that continuous Intravenous ketamine for inpatients appears to be effective, safe, well tolerated for all groups of chronic pain.The availability of hospitalisation for chronic pain patients led us propose this treatment.These findings need confirmation through large-scale,multicenter,prospective randomized controlled trials. Multimodal approaches are crucial for treating chronic pain, combining pharmacological and non-pharmacological treatments.

**Supplementary table 1.** Pain intensity NRS missing data at day 30

|  | No missing data (76 patients) |  | Missing data (13 patients) |  | p |
| --- | --- | --- | --- | --- | --- |
|  | Number with missing date | Number (%) or mean (SD) | Number with missing date | Number (%) or mean (SD) |  |
| Sex = Female | 0 | 67 (88,2%) | 0 | 11 (84,6%) | 0,66 * |
| Age | 0 | 51,66 (9,43) | 0 | 55,38 (10,57) | 0,24 ** |
| Pain intensity (NRS) | 0 | 7,54 (1,34) | 0 | 7,54 (1,45) | 0,95 ** |
| Aetiology = fibromyalgia | 0 | 49 (68,1%) | 1 | 4 (36,4%) | 0,09 * |
| HAD scale : anxiety score | 6 | 11,07 (4,79) | 0 | 12,85 (4,08) | 0,25 ** |
| HAD scale : depression score | 5 | 9,58 (4,54) | 0 | 12,31 (3,99) | 0,04 ** |
| SF 12 Mental health dimension score | 6 | 31,40 (7,96) | 1 | 31,49 (8,17) | 0,96 ** |
| SF 12 Physical health dimension score | 6 | 29,84 (7,02) | 1 | 32,29 (6,57) | 0,23 ** |
| NPSI total score | 15 | 54,72 (19,07) | 1 | 50,75 (15,18) | 0,59 ** |
| Improvement from PGIC | 0 | 52 (68,4%) | 9 | 3 (75%) | > 0,9 * |

HAD Hospital Anxiety and Depression scale ; NPSI Neuropathic Pain Symptom Inventory ; NRS Numeric rating scale ; PGIC Patient Global Impression of Change ; SF12 Short Form Health Survey
\* comparison by Fisher's exact test
\*\* comparison by Wilcoxon Rank Sum Test

## Data Availability

All data produced in the present work are contained in the manuscript

## References

1. Goldberg DS, McGee SJ. Pain as a global public health priority. BMC Public Health 2011 Oct 06;11(1):770

2. Katz, N. (2002). The impact of pain management on quality of life. Journal of pain and symptom management, 24(1), S38–S47.

3. Reid, M. C., Eccleston, C., & Pillemer, K. (2015). Management of chronic pain in older adults. BMJ 2015;350:h532

4. Cohen, Steven P., Lene Vase, and William M. Hooten. “Chronic pain: an update on burden, best practices, and new advances.” The Lancet 397.10289 (2021): 2082–2097.

5. Schwartzman, Robert J., et al. “Neuropathic central pain: epidemiology, etiology, and treatment options.” Archives of Neurology 58.10 (2001): 1547–1550.

6. Zhou, Qiang, and Morgan Sheng. “NMDA receptors in nervous system diseases.” Neuropharmacology 74 (2013): 69–75.

7. Michelet, D., et al. “Ketamine for chronic non-cancer pain: a meta-analysis and trial sequential analysis of randomized controlled trials.” European Journal of Pain 22.4 (2018): 632–646.

8. Pickering, Gisèle, et al. “Ketamine and magnesium for refractory neuropathic pain: a randomized, double-blind, crossover trial.” Anesthesiology 133.1 (2020): 154–164.

9. Cohen, Steven P., et al. “Consensus guidelines on the use of intravenous ketamine infusions for chronic pain from the American Society of Regional Anesthesia and Pain Medicine, the American Academy of Pain Medicine, and the American Society of Anesthesiologists.” Regional Anesthesia & Pain Medicine 43.5 (2018): 521–546.

10. Corriger A, Voute M, Lambert C, Pereira B, Pickering G; OKAPI Consortium. Ketamine for refractory chronic pain: a 1-year follow-up study. Pain. 2022 Apr 1;163(4):690–701.

11. Morel, V., and G. Pickering. “Kétamine et douleur chronique en France : de la théorie à la pratique.” Douleur et Analgésie 34.1 (2021): 22.

12. Pickering, Gisèle, Morel M, and Joelle Micallef. “Ketamine and chronic pain: a narrative review of its efficacy and its adverse events.” Thérapie 73.6 (2018): 529–539.

13. Voute, Marion, et al. “Ketamine in chronic pain: A Delphi survey.” European Journal of Pain 26.4 (2022): 873–887.

14. Sachau, Sendel M, Péchard M et al; Patient Reported Outcome Measures in Chronic Neuropathic Pain Clinical Trials – A Systematic Literature Review, The Journal of Pain, Volume 24, Issue 1, 2023, Pages 38–54, ISSN 1526-5900.

15. Noppers I, Niesters M, Aarts L, Smith T, Sarton E, Dahan A. Ketamine for the treatment of chronic non-cancer pain. Expert Opin Pharmacother 2010 ; 11 : 2417–2429.

16. Sigtermans, M. J., Van Hilten, J. J., Bauer et al; Ketamine produces effective and long-term pain relief in patients with Complex Regional Pain Syndrome Type 1. Pain, (2009). 145(3), 304–311.

17. Mangnus, T. J., Bharwani, K. D., Stronks, D. L., Dirckx, M., & Huygen, F. J. (2022). Ketamine therapy for chronic pain in The Netherlands: a nationwide survey. Scandinavian Journal of Pain, 22(1), 97–105.

18. Vujovic, KR Savic, et al. “A synergistic interaction between magnesium sulphate and ketamine on the inhibition of acute nociception in rats.” European Review for Medical & Pharmacological Sciences 19.13 (2015).

19. Orhurhu, Vwaire MD, MPH Orhurhu*, Mariam Salisu MD, MPH†; Bhatia, Anuj MD, Frcpc‡; Cohen, Steven P. Md§, 11. Ketamine Infusions for Chronic Pain: A Systematic Review and Meta-analysis of Randomized Controlled Trials. Anesthesia & Analgesia 129(1):p 241–254, July 2019.

20. Amr YM. Multi-day low dose ketamine infusion as adjuvant to oral gabapentin in spinal cord injury related chronic pain: a prospective, randomized, double blind trial. Pain Physician 2010; 13: 245–249

21. De Kock, M., Loix, S. and Lavand’homme, P. Ketamine and Peripheral Inflammation. CNS Neurosci Ther, (2013), 19:403–410

22. karunarathna, Indunil & Kusumarathna, et al.. Ketamine: Mechanisms, Applications, and Future Directions. (2024)

